# Psychiatric Manifestations and Associated Risk Factors among Hospitalized Patients with COVID-19 in Edo State, Nigeria

**DOI:** 10.1101/2021.10.12.21264913

**Authors:** Esther O. Okogbenin, Omonefe J. Seb-Akahomen, Osahogie I. Edeawe, Mary Ehimigbai, Helen Eboreime, Angela Odike, Michael O. Obagaye, Benjamin E. Aweh, Paul Erohubie, Williams O. Eriyo, Chinwe F. Inogbo, Peter Akhideno, Gloria Eifediyi, Reuben Eifediyi, Danny Asogun, Sylvanus A. Okogbenin

## Abstract

**Objective:** The Coronavirus Disease 2019 (COVID-19) has had devastating effects globally. These effects are likely to result in mental health problems at different levels. Although studies have reported the mental health burden of the pandemic on the general population and frontline health workers, the impact of the disease on the mental health of patients in COVID-19 treatment and isolation centres have been understudied in Africa. We estimated the prevalence of depression and anxiety and associated risk factors in hospitalized persons with COVID-19.

**Methods:** A cross-sectional survey was conducted among 489 patients with COVID-19 at the three government designated treatment and isolation centres in Edo State, Nigeria. The 9-item Patient Health Questionnaire (PHQ-9) and the Generalized Anxiety Disorder-7 (GAD-7) tool were used to assess depression and anxiety respectively. Binary logistic regression was applied to determine risk factors of depression and anxiety.

**Results:** Of the 489 participants, 49.1% and 38.0% had depressive and anxiety symptoms respectively. The prevalence of depression, anxiety, and combination of both were 16.2%, 12.9% and 9.0% respectively. Moderate-severe symptoms of COVID-19, ≥14 days in isolation, worrying about the outcome of infection and stigma increased the risk of having depression and anxiety. Additionally, being separated/divorced increased the risk of having depression and having comorbidity increased the risk of having anxiety.

**Conclusion:** A substantial proportion of our participants experienced depression, anxiety and a combination of both especially in those who had the risk factors we identified. The findings underscore the need to address these risk factors early in the course of the disease and integrate mental health interventions into COVID-19 management guidelines.

## INTRODUCTION

In recent times, the world has been challenged with a new coronavirus disease, which has demonstrated startling levels of spread, severity, and fatality all over the globe.[1] As part of the World Health Organization (WHO) response, the virus was named COVID-19 (Coronavirus Disease, 2019) and declared a pandemic.[2] Nigeria reported and hospitalized its first case of COVID-19 on 27^th^ February 2020 and has since then witnessed a steady increase in the number of cases and associated deaths.[3] Expectedly, diverse reactions to this situation have ensued, with most persons predominantly experiencing feelings of anxiety and depression about a disease of this nature and its possible outcomes.[4] The existing literature on the mental health consequences of the pandemics both on the general population and persons with confirmed COVID-19 shows that depression, anxiety, and other mental health problems are common.[5-7] These mental health consequences are predictable in both the short and long term.[8] The SARS outbreak, for instance, was accompanied by significant mental health morbidity in both patients and healthcare providers.[9]

Precisely, persons who test positive for COVID-19 or are suspected to have COVID-19 are especially vulnerable to psychological distress and mental illnesses.[8,10] Such persons may experience fear of the possible outcomes considering the potentially fatal nature of the infection. Also, staying in an isolation ward may trigger a wide range of emotions, which may include feelings of frustration, despair, hopelessness, stigma/humiliation, fear, anger and so on. Furthermore, research has demonstrated that the presence of high concentrations of inflammatory mediators above the physiologically relevant range may trigger mental disorders.[11]

The literature establishes a fundamental link between mental health and physical health. Poor physical health can lead to an increased risk of having mental health problems, conversely, poor mental health can negatively impact physical health, leading to an increased risk of some medical conditions.[12,13] Consequently, the co-existence of psychological and other medical conditions can result in increased distress, longer illness duration, poorer health outcomes and an increased cost/burden on the already scarce health resources and health care system.[8,12,13]

There is a dearth of literature on the mental health effects of COVID-19 among infected persons in Africa including Nigeria the most populous nation in Africa.[14] To the best of our knowledge, there is only one study in Nigeria that has reported the psychosocial health effects of COVID-19 infection on patients in treatment centres since the onset of the pandemic.[6] More studies are needed to understand the mental health sequelae of COVID-19 infection. Therefore, this study aimed to determine the prevalence of depression and anxiety, as well as associated risk factors in persons with COVID-19 hospitalized at the Edo state-designated treatment and isolation centres over 30 weeks.

## METHODS

### Setting and study design

A descriptive cross-sectional study was conducted from 15^th^ April to 11^th^ November 2020. The participants were COVID-19 Real Time-Reverse Transcriptase-Polymerase Chain Reaction (rRT-PCR) positive persons who were hospitalized at the three government-designated treatment and isolation centres in Edo State. These were the Irrua Specialist Teaching Hospital, Irrua, Stella Obasanjo Treatment and Isolation Centre and the University of Benin Teaching Hospital, Benin-City, Edo State, Nigeria. All three centres were government-funded.

### Participants and procedure

All eligible and consenting persons who were COVID-19 rRT-PCR positive and hospitalized at any of the study institutions within the period of the survey were recruited. The inclusion criteria comprised of persons with confirmed COVID-19, hospitalized at any of the study institutions who consented to participate in the study and were eleven years and above.

Exclusion criteria comprised of hospitalized persons who tested positive for COVID-19 but declined or were unable to give consent to participate in the study and persons below 11 years due to the inappropriateness of the assessment tools for anxiety and depression in this age group. A total of 796 patients with COVID-19 were hospitalized at the three government designated treatment and isolation centres in Edo State over the study period. Nineteen of them were below 11 years and were excluded, and 265 patients did not give consent to participate in the study. A total of 512 were recruited for the study as shown in the flowchart in Figure 1. Semi-structured and structured questionnaires incorporating socio-demographics, basic clinical history/information and an assessment of anxiety and depression were administered to recruited participants on the fifth day of admission into treatment and isolation centres.

**Figure 1:**
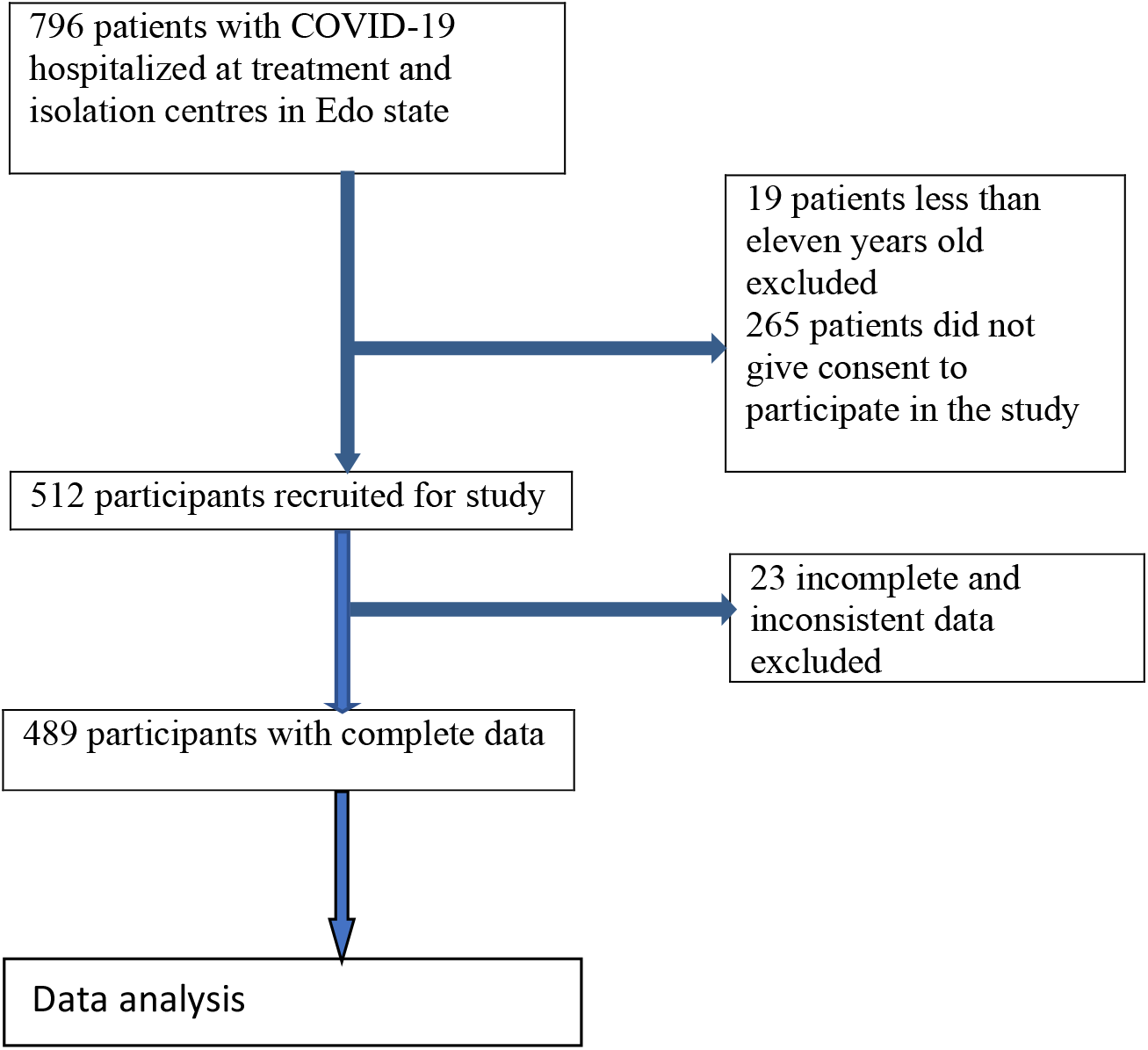
Flowchart of participants recruitment

### The socio-demographic/clinical history questionnaire

This was designed to provide information about the participant’s age, gender, marital status, employment status and the highest level of formal education. Clinical variables such as COVID-19 rRT-PCR status, previous/family history of mental illness, the severity of COVID-19 infection, the number of days in isolation, comorbidity were ascertained as well. To ascertain the worry factor, the question “what is your greatest worry about being COVID-19 positive” was asked.

### The 9-item Patient Health Questionnaire (PHQ-9)

This consists of nine items, each of which is scored 0 to 3, providing a 0 to 27 severity score.[15] PHQ-9 severity is calculated by assigning scores of 0, 1, 2, and 3, to the response categories of: Not at all, several days, more than half the days, and nearly every day, respectively. It consists of the nine criteria for depression from the Diagnostic and Statistical Manual of Mental Disorders, fourth edition (DSM-IV). The PHQ-9 is comparable or superior in operating characteristics, and valid as both a diagnostic and severity measure.[16] Scores of 5, 10, 15, and 20 represent cut-off points for mild, moderate, moderately severe, and severe depression respectively. A PHQ-9 score of 10 or greater is recommended if a single screening cut-off is to be used, this cut-off point has a sensitivity for major depression of 88% and a specificity of 88%. The modified version for adolescents PHQ-A was used for participants within the ages of 11 and 17 years. A cut-off score of ≥ 10 was used to represent cases of depression.

### The Generalized Anxiety Disorder-7 (GAD-7)

This is a 7-item self-report questionnaire that allows for the rapid detection of GAD.[21] Participants are asked if they were bothered by anxiety-related problems over the past two weeks by answering seven items on a 4-point scale. The total scores range from 0 to 21. At a cut-off score of 10, the GAD-7 had a sensitivity of 89 % and a specificity of 82 % for detecting GAD compared with a structured psychiatric interview.[17] Notably, among clinical and general population samples, the GAD-7 has demonstrated good reliability and cross-cultural validity as a measure of GAD (16). Its use has been validated in adolescents.[18] A cut-off score of ≥ 10 was used to represent cases of anxiety.

### Ethics

Ethical clearance was obtained from our institutional ethical committee. Informed written consent was obtained from each participant and from the parents or guardians of participants who were less than 18 years. Participants who were less than 18 years also assented to the study. Confidentiality and anonymity were ensured by not indicating the names of the participants on the questionnaires.

### Statistical analysis

The collected data were analysed using the Statistical Package for Social Sciences (SPSS) version 21. Dependent variables were depression and anxiety. Independent variables were sociodemographic and clinical characteristics. Descriptive statistics were used to summarise socio-demographic and clinical related data and mean with standard deviation for continuous variables. Chi-square (χ2) tests were used to test the association of independent variables with dependent variables. Fisher’s exact test was used for cells with expected frequencies < 5. The student’s *t-*test was used to compare means. Binary logistic regression was applied to identify predictors of depression and anxiety that were significant at bivariate analysis. All tests were 2-tailed, and the level of significance was set at a P-value of <0.05.

## RESULTS

### Demographics and clinical characteristics

A total of 512 patients participated in the study over the survey period. Twenty-three questionnaires were excluded from analysis due to inconsistencies and incomplete responses giving a total sample of 489 participants and a response rate of 95.5%. The mean age of participants was 43.39 (SD=16.94). Most of the participants were Christians (80.8%), married (59.3%) and slightly more than half had tertiary education (50.7%), and were employed (51.5%). Two hundred and ninety-eight (60.9%) patients had mild COVID-19 symptoms while 191 (39.1%) had moderate to severe symptoms. Of the 191 patients who had moderate to severe symptoms, 106 (55.5%) were male patients while 85 (45.5) were female patients. One hundred and seventy patients (34.8%) had comorbidity. Of the 170 (34.8) patients who had comorbidity, 69 (40.6%) had hypertension only, 33 (19.4%) had diabetes only, 31 (18.2%) had coexisting hypertension and diabetes, 9 (5.3%) were asthmatic, 3 (1.8%) had cancers and 26 (15.3%) had other illnesses. Twelve patients (2.5%) reported a history of mental illness and 12 (2.5%) also reported a family history of mental illness. Most of the patients 370 (75.7%) stayed less than 14 days in isolation while 119 (24.3%) stayed 14 or more days in isolation. Demographics and clinical characteristics of participants are reported in Table 1.

**Table 1:**
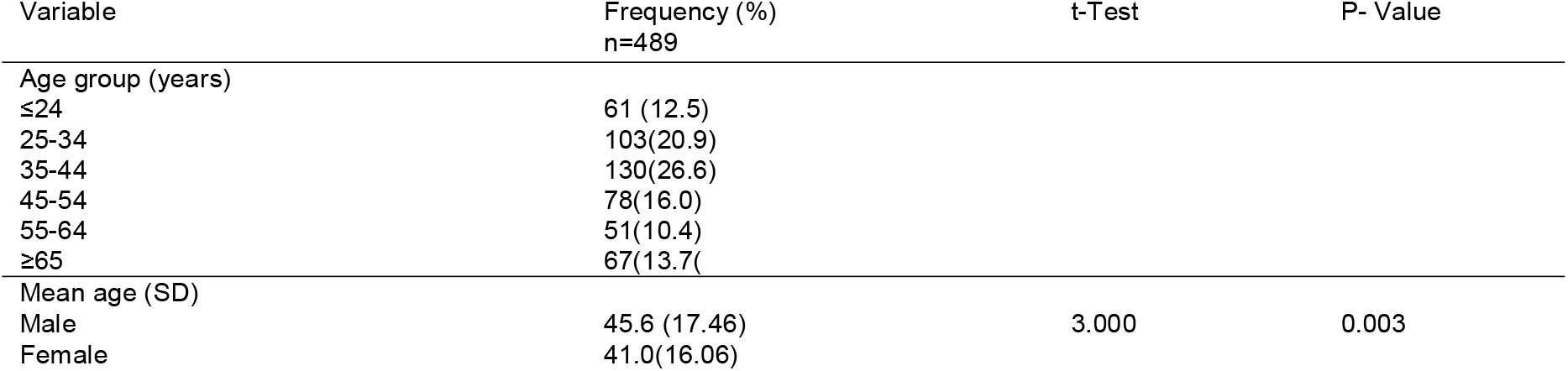
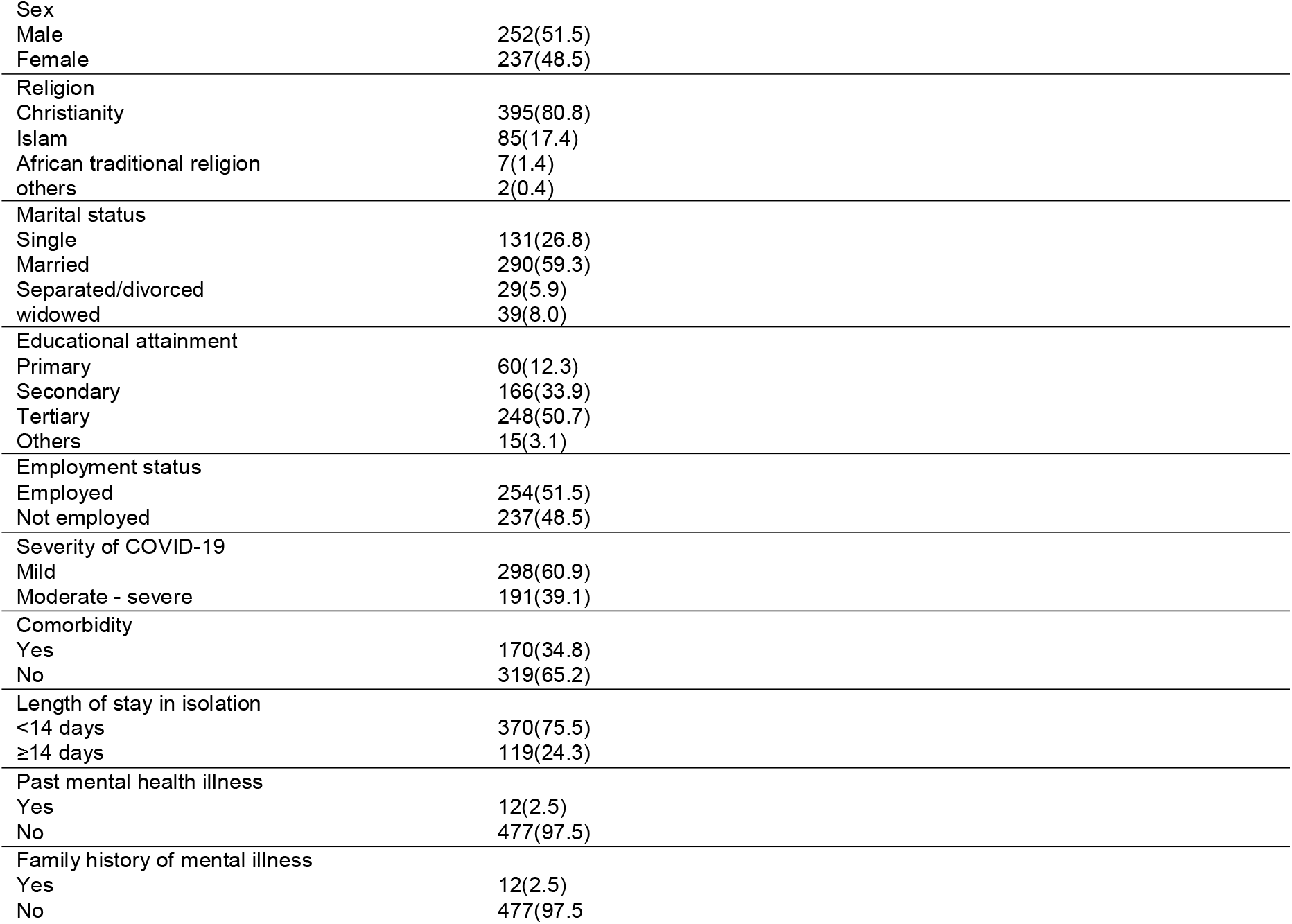
Demographic and clinical characteristics of hospitalized patients with COVID-19 in Edo State, Nigeria

| Variable | Frequency (%)<br>n=489 | t-Test | P- Value |
| --- | --- | --- | --- |
| Age group (years) |  |  |  |
| ≤24 | 61 (12.5) |  |  |
| 25-34 | 103(20.9) |  |  |
| 35-44 | 130(26.6) |  |  |
| 45-54 | 78(16.0) |  |  |
| 55-64 | 51(10.4) |  |  |
| ≥65 | 67(13.7) |  |  |
| Mean age (SD) |  |  |  |
| Male | 45.6 (17.46) | 3.000 | 0.003 |
| Female | 41.0(16.06) |  |  |

|  |  |
| --- | --- |
| Sex |  |
| Male | 252(51.5) |
| Female | 237(48.5) |
| Religion |  |
| Christianity | 395(80.8) |
| Islam | 85(17.4) |
| African traditional religion | 7(1.4) |
| others | 2(0.4) |
| Marital status |  |
| Single | 131(26.8) |
| Married | 290(59.3) |
| Separated/divorced | 29(5.9) |
| widowed | 39(8.0) |
| Educational attainment |  |
| Primary | 60(12.3) |
| Secondary | 166(33.9) |
| Tertiary | 248(50.7) |
| Others | 15(3.1) |
| Employment status |  |
| Employed | 254(51.5) |
| Not employed | 237(48.5) |
| Severity of COVID-19 |  |
| Mild | 298(60.9) |
| Moderate - severe | 191(39.1) |
| Comorbidity |  |
| Yes | 170(34.8) |
| No | 319(65.2) |
| Length of stay in isolation |  |
| <14 days | 370(75.5) |
| ≥14 days | 119(24.3) |
| Past mental health illness |  |
| Yes | 12(2.5) |
| No | 477(97.5) |
| Family history of mental illness |  |
| Yes | 12(2.5) |
| No | 477(97.5) |

### Prevalence of depression and anxiety

About half of the patients 240 (49.1%) had mild to severe depressive symptoms, 161 (32.9%) had mild symptoms, 62 (12.7) had moderate symptoms, 12 (2.5%) had moderately severe and 5 (1.0%) had severe symptoms of depression. More than a third of the patients 186 (38.0%) had mild to severe anxiety symptoms, 124 (25.4%) had mild symptoms, 47 (9.6) had moderate symptoms, and 15 (3.1%) had severe symptoms of anxiety. Seventy-nine (16.2%) patients were classified as cases of depression while 63 (12.9%) patients were classified as cases of anxiety and 44 (9.0%) as cases of anxiety comorbid with depression. Prevalence of depression and anxiety are reported in Table 2.

**Table 2:**
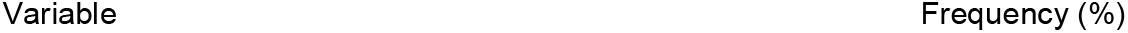
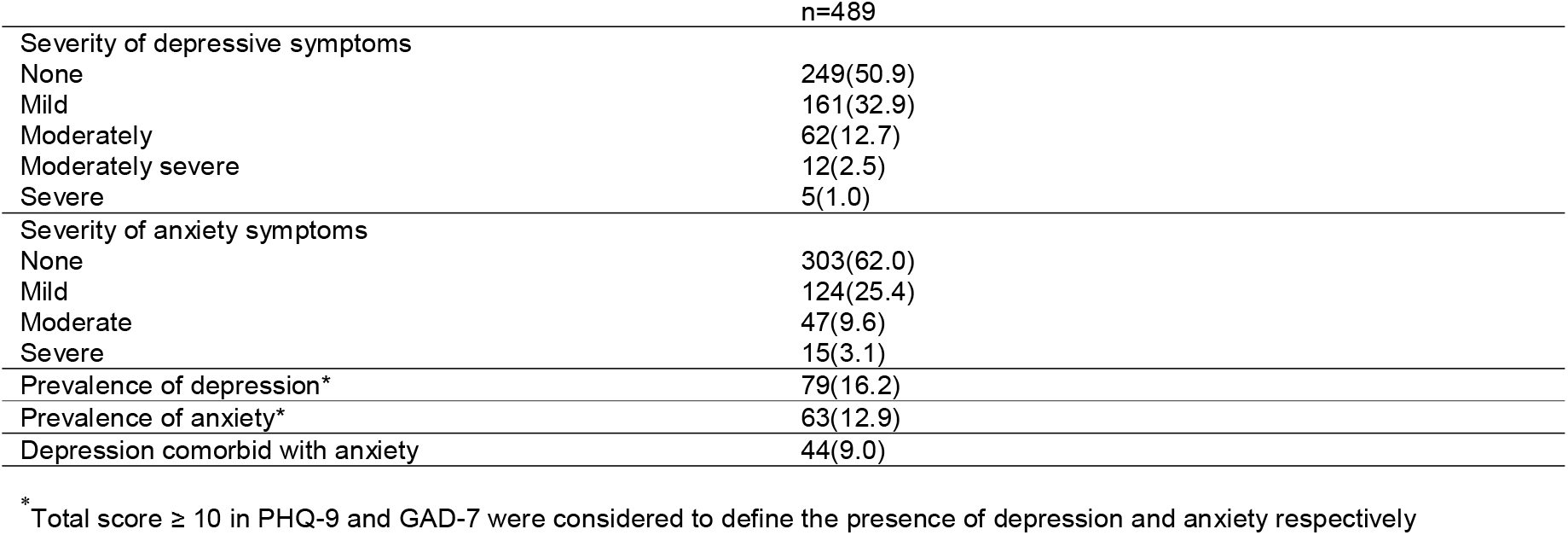
Prevalence of depression and anxiety among hospitalized patients with COVID-19 in Edo State, Nigeria

|  | n=489 |
| --- | --- |
| Severity of depressive symptoms |  |
| None | 249(50.9) |
| Mild | 161(32.9) |
| Moderately | 62(12.7) |
| Moderately severe | 12(2.5) |
| Severe | 5(1.0) |
| Severity of anxiety symptoms |  |
| None | 303(62.0) |
| Mild | 124(25.4) |
| Moderate | 47(9.6) |
| Severe | 15(3.1) |
| Prevalence of depression* | 79(16.2) |
| Prevalence of anxiety* | 63(12.9) |
| Depression comorbid with anxiety | 44(9.0) |

\* Total score $\geq 10$ in PHQ-9 and GAD-7 were considered to define the presence of depression and anxiety respectively

### Worry factors

Ninety (18.4%) patients reported no worry about being COVID-19 positive. The most frequent worry factor expressed was worrying about the outcome of illness 195 (39.9%). Worry factors are reported in Figure 2.

**Figure 2:**
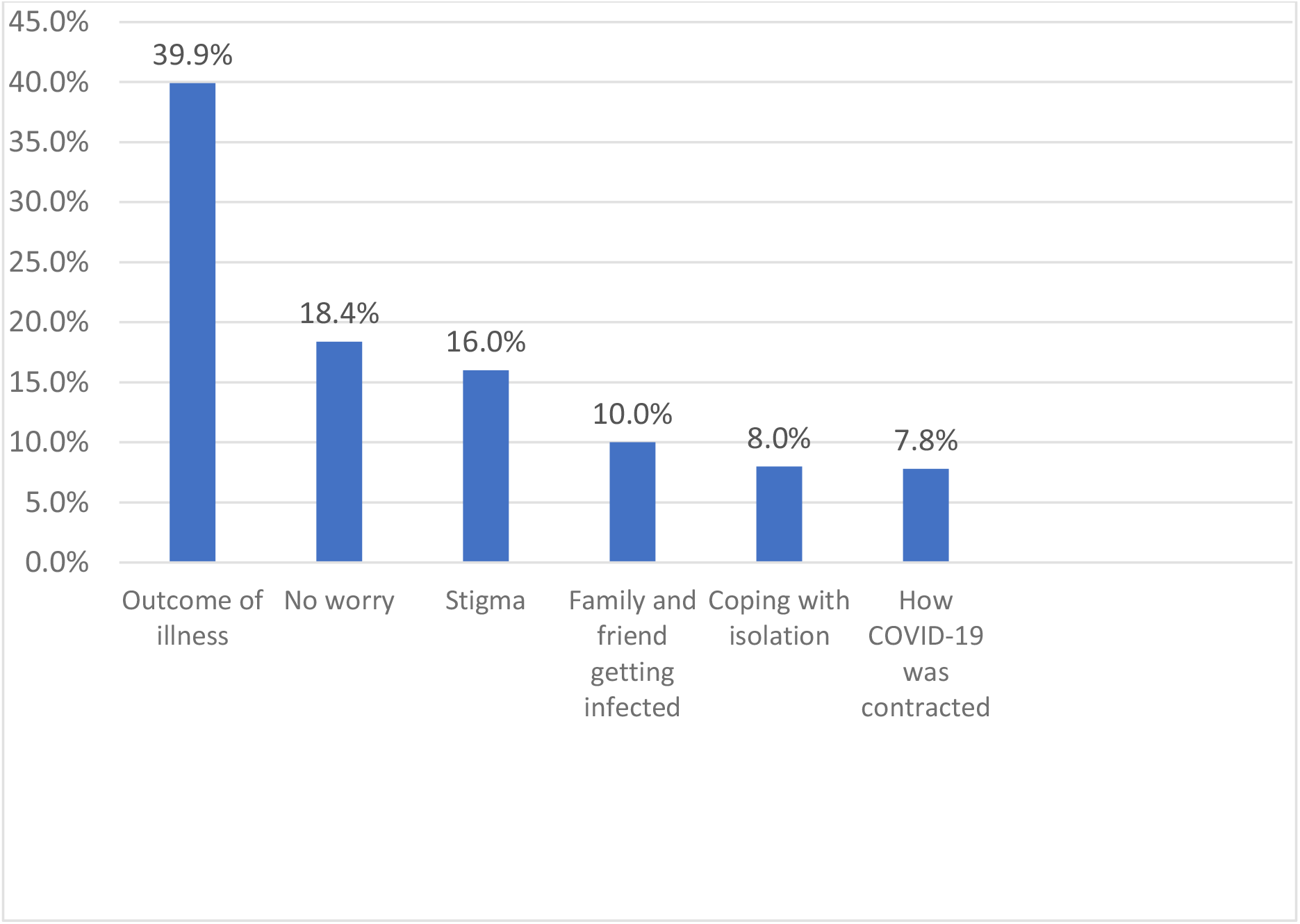
Worry factors expressed by hospitalized patients with COVID-19 in Edo State, Nigeria

### Risk factors associated with depression and anxiety

Using bivariate analysis, being separated/widowed (χ^2^ = 10.943, P =0.012), having moderate to severe COVID-19 symptoms (χ^2^ = 5.302, P = 0.021), staying in isolation for 14 days or more (χ^2^ = 11.368, P = 0.001) and having outcome of illness as worry factor (18.056, P = 0.002) were found to be significantly associated with cases of depression. Having moderate to severe COVID-19 symptoms (χ^2^ = 5.302, P = 0.021), staying in isolation for 14 days or more (χ^2^ = 11.368, P = 0.001), having outcome of illness as worry factor (18.056, P = 0.002) and having a comorbidity were found to be significantly associated with cases of anxiety.

Analysis of factors associated with depression and anxiety are shown in Table 3.

**Table 3:**
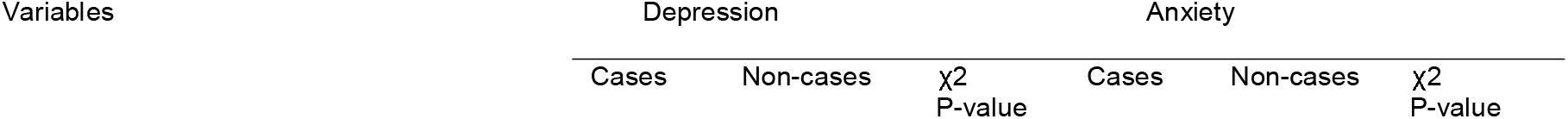
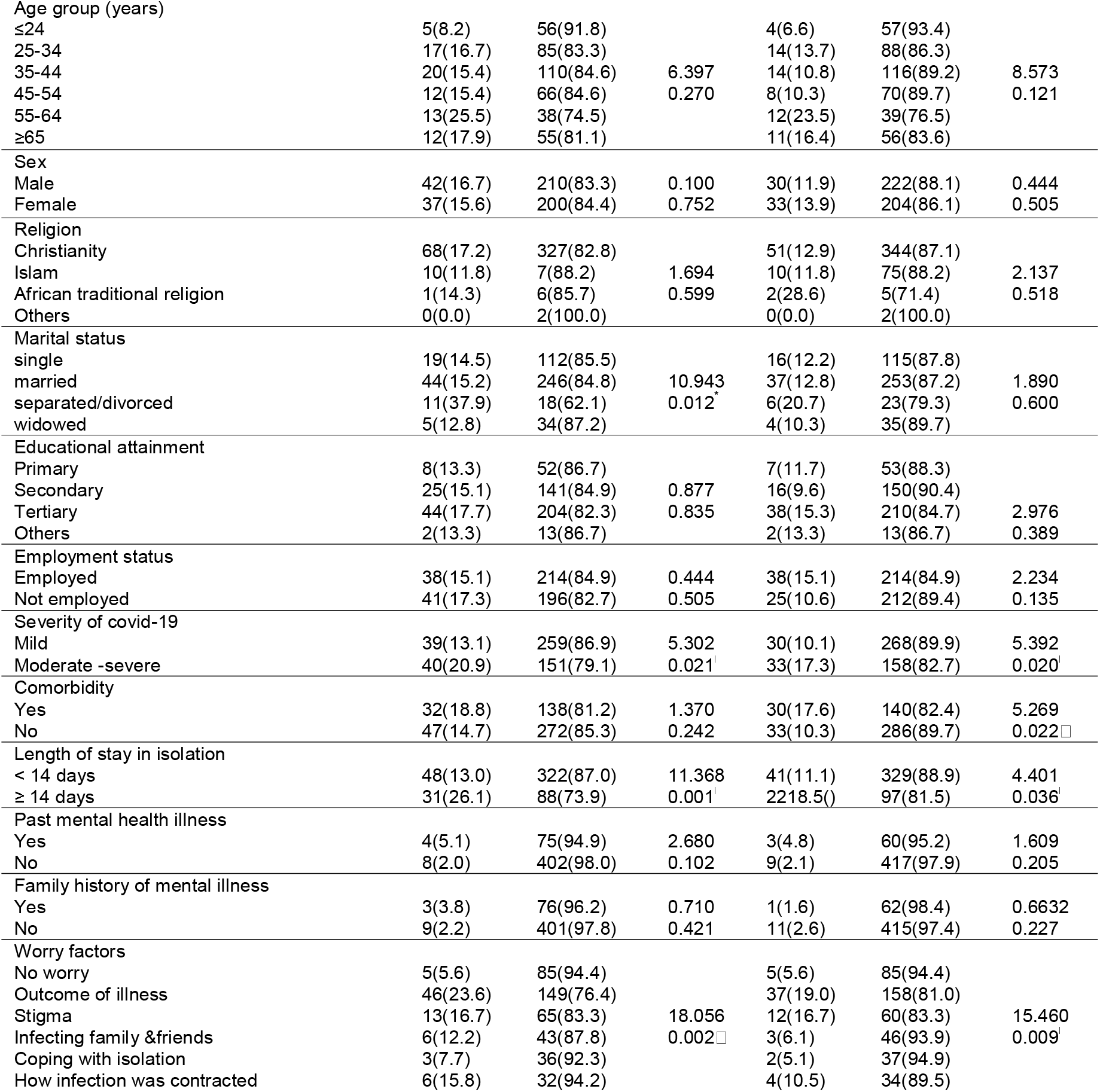
Association of demographic and clinical characteristics with depression and anxiety among hospitalized patients with COVID-19 in Edo State, Nigeria

### Predictors of depression and anxiety

We conducted a binary logistic regression analysis to measure the correlations between dependent and independent variables that were significant at bivariate analysis. The unadjusted odds ratios are displayed in Table 4.

**Table 4:** Logistic regression (with unadjusted odds ratios) of demographic and clinical risk factors associated with depression and anxiety among hospitalized patients with COVID-19 in Edo State, Nigeria

| Variables | Depression |  |  | Anxiety |  |  |
| --- | --- | --- | --- | --- | --- | --- |
|  | OR | P value | 95% CI | OR | P value | 95% CI |
| Marital status |  |  |  |  |  |  |
| Married | 1(Ref) |  |  | - |  |  |
| Single | 0.95 | 0.859 | 0.530 – 1.698 | - | - | - |
| Separated/divorced | 3.42 | 0.003* | 1.511 – 7.725 | - | - | - |
| widowed | 0.82 | 0.699 | 0.305 – 2.217 | - | - | - |
| Severity of COVID-19 |  |  |  |  |  |  |
| Mild | 1(Ref) |  |  | 1(Ref) |  |  |
| Moderate -Severe | 1.76 | 0.022* | 1.084 - 2.856 | 1.87 | 0.022* | 1.096 – 3.176 |
| Duration of stay in isolation |  |  |  |  |  |  |
| < 14 days | 1(Ref) |  |  | 1(Ref) |  |  |
| ≥ 14 days | 2.36 | 0.001* | 1.420 - 3.933 | 1.82 | 0.038* | 1.034 – 3.203 |
| Worry factor |  |  |  |  |  |  |
| No worry | 1(Ref) |  |  | 1(Ref) |  |  |
| Outcome of illness | 5.25 | 0.001* | 2.008 – 13.715 | 3.98 | 0.005* | 1.509 – 10.506 |
| Stigma | 3.40 | 0.026* | 1.154 – 10.020 | 3.09 | 0.043* | 0.037 – 9.209 |
| Infecting family & friends | 2.37 | 0.173 | 0.685 – 1.215 | 1.11 | 0.891 | 0.253 – 4.849 |
| Coping with isolation | 1.42 | 0.645 | 0.321 – 6.245 | 0.92 | 0.922 | 0.170 – 4.953 |
| How COVID-19 was contacted | 3.19 | 0.070 | 0.909 – 11.176 | 2.00 | 0.323 | 0.506 – 7.899 |
| Comorbidity |  |  |  |  |  |  |
| No | - | - | - | 1(ref) |  |  |
| Yes | - | - | - | 1.86 | 0.023* | 1.089 – 3.168 |

Analysis for adjusted odds ratios was conducted to control for confounders. The model explained 14.1% (Nagelkerke *R*^*2*^) of the variance in depression and correctly classified 83.6% of cases. Being separated/divorced (AOR = 3.98, P = 0.002, 95% CI =1.655 - 9.583) compared to being married, staying 14 days or longer in isolation (AOR =2.31, P = 0.002, 95% CI = 1.352 - 3.946) compared to staying less than 14 days and worrying about the outcome of COVID-19 infection (AOR = 4.98, P = 0.002, 95% CI = 1.833 - 13.506) compared to no worry significantly increased the odds of having depression. For anxiety, the logistic model explained 8.2% (Nagelkerke *R*^*2*^) of the variance and correctly classified 87.1% of cases. Only worrying about the outcome of COVID-19 infection (AOR = 3.23, P = 0.023, 95% CI = 1.176 – 8.155) significantly increased the odds of cases of anxiety after controlling for confounding variables.

## DISCUSSION

We present the estimates of depression and anxiety and associated risk factors from a cross-sectional cohort of hospitalised patients with confirmed COVID-19 in the three treatment and isolation centres in Edo State, Nigeria. Our study shows that male patients were slightly more than female patients. Fadipe et al also found a preponderance of male patients in confirmed COVID-19 cases.[6] The disparity between males and females may be attributed to inequalities in health-seeking behaviour. Males may access health care facilities and testing more than females.[19] However, Peckham et al had demonstrated no significant difference in the proportion of males and females with confirmed COVID-19 infection in a meta-analysis of over 3 million reported global cases.[20] Notably, our finding that moderate-severe form of COVID-19 infection was more in males than females was supported by Peckham et al in his meta-analysis where he demonstrated that male patients had significant higher odds of intensive care unit admissions and deaths than female patients and concluded that males and females were at equivalent risk of COVID-19 infection and male sex was associated with the development of severe disease.[20] This increased severity may be why males present more than females to health facilities for testing and treatment.

We found high rates of symptoms of depression and anxiety in patients with COVID-19 in treatment and isolation centres. About half (49.1%) and more than a third (38.0%) of the patients had symptoms of depression and anxiety respectively. Similarly, high rates of depressive and anxiety symptoms were reported by Paz et al in Ecuador and Zhang et al in China in patients with COVID-19.[7,21] As the pandemic spreads rapidly around the world with devastating effects, it is expected that an increasing number of persons would experience anxiety, depression, and other mental health problems especially in the context of a confirmed infection. This is made worse by the inappropriate risk communication and misinformation on social media.[5] Although in this study, apparent lower prevalence rates were recorded for patients classified as cases of depression (16.2%), anxiety (12.7%) and a combination of both (9.0%) when compared to the proportion having symptoms, these rates are higher than the prevalence rates of depression (5.5%), anxiety (3.5%) and anxiety comorbid with depression (1.2%) reported in the general population in the pre-COVID-19 pandemic era in Lagos, Nigeria.[22] A similar study in patients with COVID-19 in treatment centres in Lagos, Nigeria, reported higher rates for probable cases of depression (28.10%), anxiety (27.50%) and a combination of both (15.60%).[6] However, the study conducted in Lagos, Nigeria, had a smaller number of participants compared to ours (160 versus 489) and was conducted early in the first wave of the pandemic which coincided with peaks in COVID-19 deaths. Additionally, Lagos is the epicentre with the highest number of deaths from the pandemic in Nigeria.[3]. These factors may be responsible for the higher rates of depression and anxiety they reported. On the other hand, we found higher rates of cases of depression and anxiety than those reported in recovered patients with COVID-19 in Tehran, Iran.[23]

The high rates of depressive and anxiety symptoms found in this study suggest that a substantial proportion of hospitalized patients with COVID-19 suffer the additional distress of concurrent mental health symptoms/conditions. While the development of depression and anxiety may partly result from the psychosocial consequences of COVID-19, they may also be induced by direct neurological injury through hypoxic damage and neuro-invasion.[24] COVID-19 disease has been described as a cytokine release syndrome with increased serum concentrations of interleukin-6 and other inflammatory cytokines which have been associated with psychiatric manifestations.[11,24] These manifestations may not only persist beyond the pandemic but may lead to increased illness, prolonged stay in the hospital, poorer health outcomes and avoidable strain on health care systems and scarce resources if not recognised and managed promptly.[12,13] Okogbenin et al in a retrospective analysis of psychiatric consultations in patients with Lassa fever infection concluded that mental health intervention could improve overall outcomes of Lassa fever disease.[25] Also, Okogbenin and Seb-Akahomen had recommended mental health and psychosocial intervention via tele mental health services as a useful alternative to face-to-face intervention, a model they reported helped ameliorate the mental health effects of COVID-19 infection in patients at the Irrua Specialist Teaching Hospital’s treatment and isolation centre in Nigeria.[26] It is becoming apparent that deliberate efforts are needed to address the mental health and psychosocial effects of COVID-19 in persons with the disease.

Being separated/divorced was a strong predictor of depression compared to being married in our study. This is in tandem with previous studies that reported a significant association between being separated/divorced and experiencing depression.[22,27] We note that depression and anxiety occurred most in patients aged 55 years and above and least in those who were 24 years or less. Similarly, Zhang et al and Kong et al in China reported that older patients with COVID-19 were more likely to have depression and anxiety.[21,28] Older persons are generally more likely to have comorbidity that have been associated with a severe form of COVID-19.[29] In the analysis of the severity of COVID-19, we found that having moderate to severe symptoms significantly increased the risk of having depression and anxiety. This may occur for two reasons. Firstly, they may have a more intense immune-inflammatory dysregulation.[30] and secondly, the awareness of the relationship between the severity of COVID-19 and its outcomes in these patients may precipitate depression and anxiety. This may also explain why patients with comorbidity had an increased risk for anxiety.

Staying 14 days or longer in isolation was significantly associated with increased risk of depression and anxiety. This compares with other studies that have linked social isolation during the pandemics with depression, anxiety, and other mental health disorders.[29,30] Loss of functionality and social interaction, limited physical activities, suffering without the support of loved ones, and watching others suffer or even die are possible contributory factors that may precipitate mental health problems in COVID-19 treatment and isolation centres. Conversely, the presence of depression and/or anxiety could be a possible reason for a prolonged stay in isolation. Also, we found that most of our participants (81.6%) expressed varying worry factors. In addition, worrying about the outcome of the infection was the strongest predictor for depression and anxiety even after adjusting for covariance. Other studies have reported similar findings.[5,6] This is not unexpected considering the potential of the disease to cause a severe illness and have a fatal outcome. Worrying about Stigma also increased the risk of having depression and anxiety. Fadipe et al reported similar concerns in a comparable cohort in Lagos (6). The very stringent measures of societal lockdown, social distancing, constant hand hygiene and heavy use of personal protective equipment by COVID-19 frontline health professionals to prevent transmission of the virus may promote discrimination and stigma including self-stigma. It is therefore quite understandable for isolated and hospitalized persons with COVID-19 to be worried about stigma.

### Strengths and limitations of this study

As far as we know, this is the second study exploring psychiatric manifestations in hospitalized patients with COVID-19 in Nigeria, the most populous country in Africa. The study identified important risk factors. Also, we conducted a multi-centre study that collected samples over 30 weeks in the ongoing pandemic, this increased our sample size compared to the previous study, giving more room for generalization of findings. Our study had some limitations. Firstly, it had a cross-sectional design and so could not permit causal inferences. Secondly, we did not have a control group, this minimised our ability to estimate the true impact of COVID-19 on the mental health of our participants and thirdly, a structured diagnostic interview was not used to confirm our cases of depression and anxiety.

In conclusion, we report that a considerable proportion of hospitalized patients with COVID-19 had depression, anxiety, and a combination of both. Risk factors identified should be monitored and addressed promptly when managing patients with COVID-19. Mental health interventions and psychosocial support need to be integrated into COVID-19 management guidelines to ensure holistic care and ameliorate the mental health effects of the disease.

Prospective studies are recommended to determine the true mental health impact of COVID-19 on persons with the disease.

## Data Availability

All data produced in the present study are available upon reasonable request to the authors

## Data availability statement

Data are available from the corresponding author on reasonable request.

## Ethics statement

Patient consent for publication Not required

## Ethics approval

The study was approved by the Irrua Specialist Teaching Hospital Research Ethics Committee-Protocol No: ISTH/HREC/20202004/065.

